# Auto-segmentation of hemi-diaphragms in free-breathing dynamic MRI of pediatric subjects with thoracic insufficiency syndrome

**DOI:** 10.1101/2024.09.17.24313704

**Authors:** Yusuf Akhtar, Jayaram K. Udupa, Yubing Tong, Caiyun Wu, Tiange Liu, Leihui Tong, Mahdie Hosseini, Mostafa Al-Noury, Manali Chodvadiya, Joseph M. McDonough, Oscar H. Mayer, David M. Biko, Jason B. Anari, Patrick Cahill, Drew A. Torigian

## Abstract

**Purpose:** In respiratory disorders such as thoracic insufficiency syndrome (TIS), the quantitative study of the regional motion of the left hemi-diaphragm (LHD) and right hemi-diaphragm (RHD) can give detailed insights into the distribution and severity of the abnormalities in individual patients. Dynamic magnetic resonance imaging (dMRI) is a preferred imaging modality for capturing dynamic images of respiration since dMRI does not involve ionizing radiation and can be obtained under free-breathing conditions. Using 4D images constructed from dMRI of sagittal locations, diaphragm segmentation is an evident step for the said quantitative analysis of LHD and RHD in these 4D images.

**Methods:** In this paper, we segment the LHD and RHD in three steps: recognition of diaphragm, delineation of diaphragm, and separation of diaphragm along the mid-sagittal plane into LHD and RHD. The challenges involved in dMRI images are low resolution, motion blur, suboptimal contrast resolution, inconsistent meaning of gray-level intensities for the same object across multiple scans, and low signal-to-noise ratio. We have utilized deep learning (DL) concepts such as Path Aggregation Network and Dual Attention Network for the recognition step, Dense-Net and Residual-Net in an enhanced encoder-decoder architecture for the delineation step, and a combination of GoogleNet and Recurrent Neural Network for the identification of the mid-sagittal plane in the separation step. Due to the challenging images of TIS patients attributed to their highly distorted and variable anatomy of the thorax, in such images we localize the diaphragm using the auto-segmentations of the lungs and the thoraco-abdominal skin.

**Results:** We achieved an average±SD mean-Hausdorff distance of ∼3±3 mm for the delineation step and a positional error of ∼3±3 mm in recognizing the mid-sagittal plane in 100 3D test images of TIS patients with a different set of ∼430 3D images of TIS patients utilized for building the models for delineation, and separation. We showed that auto-segmentations of the diaphragm are indistinguishable from segmentations by experts, in images of near-normal subjects. In addition, the algorithmic identification of the mid-sagittal plane is indistinguishable from its identification by experts in images of near-normal subjects.

**Conclusions:** Motivated by applications in surgical planning for disorders such as TIS, we have shown an auto-segmentation set-up for the diaphragm in dMRI images of TIS pediatric subjects. The results are promising, showing that our system can handle the aforesaid challenges. We intend to use the auto-segmentations of the diaphragm to create the initial ground truth (GT) for newly acquired data and then refining them, to expedite the process of creating GT for diaphragm motion analysis, and to test the efficacy of our proposed method to optimize pre-treatment planning and post-operative assessment of patients with TIS and other disorders.

## 1. INTRODUCTION AND RELATED WORK

The abnormalities of respiration that occur in certain disorders like thoracic insufficiency syndrome (TIS) [1], which are linked to an abnormal architecture of the thorax including the diaphragm, can be better understood in terms of extent and severity by studying the regional structure and function of the left hemi-diaphragm (LHD) and right hemi-diaphragm (RHD) individually. This improved understanding can in turn help us to better comprehend the effects of various deformities of the spine, chest wall, and other structures upon diaphragmatic structure and function. In turn, this information can potentially serve to better determine and optimize the surgical approach to be utilized in individual patients, including for example the use of devices such as the Vertical Expandable Prosthetic Titanium Rib (VEPTR). Dynamic magnetic resonance imaging (dMRI) is an ideal imaging modality for capturing dynamic images, unlike dynamic computed tomography (CT) which involves the exposure of subjects to ionizing radiation. With dMRI, image acquisition of the thoraco-abdominal region is obtained under free-breathing conditions [2], which is a major advantage in terms of practical implementation and patient comfort, as patients often have a limited capacity to follow specific instructions for controlled breathing such as in the setting of TIS. As a preliminary step to understand what is normal and abnormal in terms of regional respiratory structure and function, we first require the acquisition of dMRI images of the thorax in near-normal subjects to create a reference model of the normal thorax. As such, this paper deals with dMRI images obtained in both near-normal subjects and TIS patients.

Utilizing the method of [2], 4D images are first constructed from the acquired dMRI images. The 4D images are then used to segment the diaphragm, allowing for further analysis of diaphragmatic structure and function. Segmentation of the diaphragm in dMRI images can be obtained either manually or with the help of automatic segmentation algorithms. The former approach is time-consuming, labor-intensive, and error-prone [3], especially considering the 4D nature of the images and the many slices that need to be contoured. In this paper, we present a method for the latter, namely automatic segmentation of the diaphragm in dMRI images.

dMRI images pose the following challenges (see Figure 1) for segmentation of the diaphragm (Dph): (1) different meanings of gray-level intensities for Dph and other objects for the same subject across different image acquisitions, and for different subjects, (2) poor contrast amongst certain objects in dMRI images and drastically varying degrees of contrast along a given boundary between objects, (3) low signal-to-noise ratio, (4) motion blur, (5) relatively low spatial resolution, (6) the highly spatially sparse nature of the Dph where Dph and its external boundary are roughly the same, and (8) the lack of an explicit intensity boundary between LHD and RHD of Dph.

**Figure 1:**
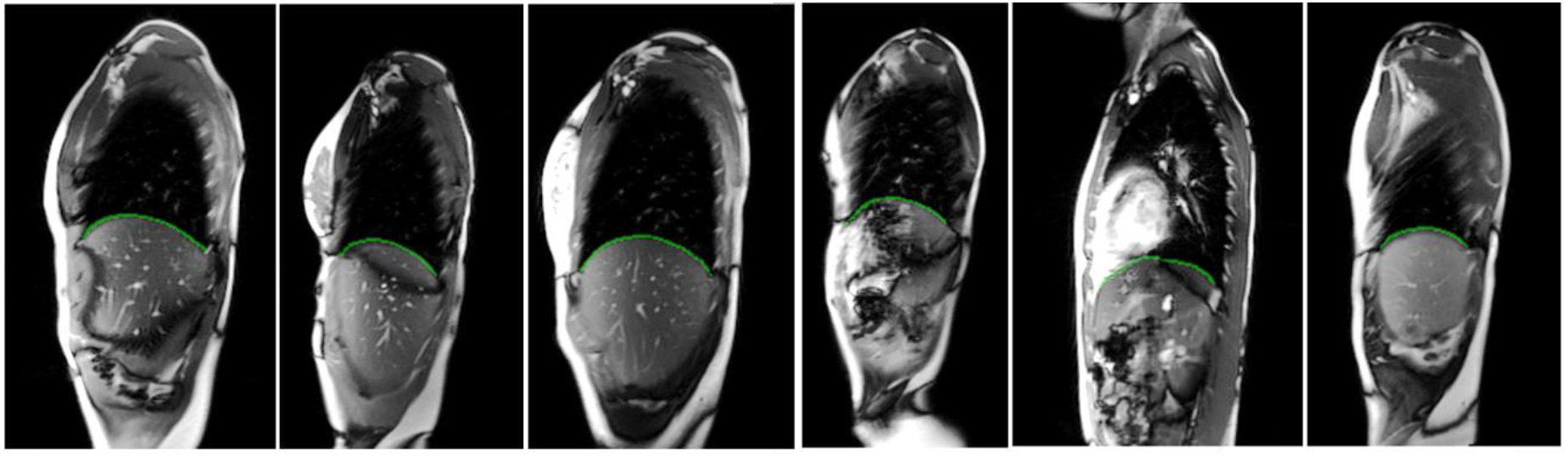
Automatic delineations (green outlines) of right hemi-diaphragm (3 images shown on the left) and left hemi-diaphragm (3 images shown on the right) on sagittal dMRI images.

In [4], the Dph was segmented in 4D CT images (voxel resolution of 0.98 to 1.29 mm in-slice and 3 mm inter-slice) of 9 cancer patients (age not disclosed) by finding the contact surfaces of lungs, heart, rib cage, and descending thoracic aorta with the Dph, reporting an average±SD of mean-Hausdorff distance (mean-*HD*) of 2.55±0.39 mm. In [5], the Dph was segmented using quadratic modeling of the contact surface of the auto-segmentation of the lungs only in 9 3D CT images (voxel resolution of 0.35 to 0.55 mm in-slice and 5 mm inter-slice) of 4 pediatric subjects (age: 2 weeks to 11 years) with neuroblastoma, producing an average mean-*HD* of 5.85 mm. In [6], auto-segmentation of the lungs was utilized to locate the Dph and chest wall in cine-MRI (slice thickness of 6 mm) of 20 young (mean age: 10.5 years) patients with muscular dystrophy, using b-spline interpolation techniques, yielding an average mean-*HD* of 1.2-1.3 mm. The auto-segmentation of the lungs using simple thresholding was utilized in [7] to locate the Dph using optimal surface selection in volumetric graphs, which produced an average mean-*HD* of 18.3 mm in CT images (voxel resolution of 0.68 mm x 0.68 mm x 3 mm) of 7 randomly selected patients (age not disclosed). A recent article [8] used the segmentation of the Dph to analyze its curvature and motion over time in MRI scans (voxel resolution of 3 mm x 3 mm x 3 mm) of 30 patients (mean age: 43 years) with Pompe disease and 10 healthy controls (mean age: 37 years). The authors of [8] did not disclose their segmentation algorithm and did not report any performance metric for segmentation of the Dph.

As evident from the aforementioned articles, the Dph has been segmented using auto-segmentation of the lungs in mostly CT images reconstructed in the axial plane. We have also used auto-segmentations of the lungs in sagittal dMRI images for segmentation of the Dph of pediatric subjects but in conjunction with inter-slice information. For example, since sagittal plane images through the thorax located in proximity to or passing through the heart often do not contain the lungs, we developed an alternative method to localize the Dph in such sagittal slices using those sagittal slice locations which do contain the lungs. Furthermore, given the challenges associated with dMRI images listed above, we cannot use the algorithms of [4, 5, 6, 7, 8] for segmentation of the lungs directly. Instead, we use two deep learning (DL) networks; one for recognition (localization using bounding boxes) of the lungs with [9] employing concepts such as Path Aggregation Networks [10] and Dual Attention Networks [11], and the other for delineation (within the bounding boxes) of the lungs with an enhanced version of an encoder-decoder architecture [12]. In contradistinction to the related works described in the previous paragraph that dealt with a few tens of images, we evaluate 100 test 3D images and utilize ∼430 3D images for training our DL models. A preliminary version [13] of our work was presented at the SPIE Medical Imaging Conference in 2024. This current paper differs from the conference paper in the following respects.

i. The conference paper presented a method for segmentation of the Dph using segmentation models for seven other objects (two lungs, two kidneys, liver, spleen, and thoraco-abdominal skin). In this paper, we segment the Dph using the auto-segmentation of the two lungs and the thoraco-abdominal skin only.
ii. The results in the conference paper were presented for near-normal subjects only. In the current paper, we evaluate the performance of our method in pediatric patients with TIS, where the images are more challenging to handle compared to images of near-normal subjects due the highly distorted and inconsistent anatomy of the thorax in TIS.
iii. The results in the conference paper were obtained with 300 images of near-normal subjects whereas the results in this paper are obtained with ∼380 images of near-normal subjects and ∼530 images of TIS patients.

## 2. METHODS

During dMRI acquisition, an image slice is first acquired continuously at one fixed sagittal location for a specified duration (typically over 10 respiratory cycles), and then the next sagittal location is dynamically imaged for the next specified duration, and so on until the image of the entire thoraco-abdominal region is fully captured from right to left. However, to segment the Dph, we first perform a 4D construction of the body region image with the intent of representing the dynamic body region by the 4D image over one respiratory cycle via an optical flux strategy [2]. Then, we segment the Dph in the 3D image corresponding to any specified respiratory phase of interest such as the end-inspiration (EI) and end-expiration (EE) time points.

We follow the paradigm of splitting the heavy-duty task of segmentation of an object of interest into two steps: *recognition* and *delineation*. In the recognition step, we localize the object of interest in the unseen image using bounding boxes. The delineation step involves marking the outline of the object within the bounding boxes. In [14], we reported methods for the segmentation of the lungs in dMRI images of healthy subjects. In that study, we showed that an artificial intelligence module called Deep Learning-Recognition (DL-R) [9] performs well for the recognition step. For the delineation step, ABCNet [12], another artificial intelligence network, performed quite well despite the challenges involved with dMRI images. We adapt these approaches, DL-R and ABCNet, as some of the modules for segmentation of the Dph in a novel pipeline in the current work.

Obstructions to motion of the hemi-diaphragms differ between right and left sides, given asymmetrical positions of adjacent organs such as the heart, different sizes and shapes of the left and right lungs, and different and asymmetrically situated organs in the abdomen. We refer to the portion of the Dph to the left of the mid-sagittal plane as the LHD, and the portion of the Dph to the right of the mid-sagittal plane as the RHD. We utilized a combination of CNN and RNN [15] to identify the sagittal slice containing the mid-sagittal plane. We take the prediction of the portion of the Dph to the left or right of this mid-sagittal slice to be LHD or RHD, respectively. In this paper, we demonstrate our system for auto-segmentation of the entire Dph, LHD, and RHD in pediatric dMRI by utilizing our extensive experience and tools we previously developed for segmenting organs body-wide in CT images [9, 12, 15].

The segmentation of LHD and RHD involves three sequential steps. These are (1) recognition of the entire Dph, (2) delineation of the entire Dph, and (3) splitting of the Dph along the mid-sagittal plane into LHD and RHD. In this section, we describe these modules. They are referred to diaphragm-recognition (Dph-R) [9], diaphragm-delineation (Dph-D) [12], and diaphragm-separation (Dph-S) [15] in subsections 2.1, 2.2, and 2.3, respectively. We now discuss details of our dataset in the next subsection.

### 2.0 Data acquisition and pre-processing

#### 1. dMRI scans

The dMRI scan data were acquired from 189 6-20-year-old healthy children and 149 pediatric TIS patients under an ongoing prospective research study protocol approved by the Institutional Review Board at the Children’s Hospital of Philadelphia (CHOP) and University of Pennsylvania, with a Health Insurance Portability and Accountability Act waiver. All participants’ or their legal representatives’ consent were taken for the present study. We excluded scans with significant body movement during scanning or with obvious image artifacts. The thoracic dMRI protocol includes a 3T MRI scanner (Verio, Siemens, Erlangen, Germany) using a True-FISP bright-blood sequence with acquisition and reconstruction parameters of TR=3.82 ms, TE=1.91 ms, flip angle 76°, bandwidth 258 Hz, 320×320 matrix, and voxel size ∼1×1×6 mm^3^. We had pre-operative (before VEPTR placement) and post-operative (after VEPTR placement) dMRI images of 49 TIS patients, pre-operative only dMRI images of 70 TIS patients, and post-operative only dMRI images of 30 TIS patients. For each sagittal location across the thorax, 80 image slices were obtained over several tidal breathing cycles at ∼480 ms/slice. On average, 35 sagittal locations across the thorax were imaged. Therefore, a total of 2800 (35 × 80) 2D MRI slices were acquired per subject.

#### 2. 4D construction

Given the dMRI scan for each subject, a small set of 175-320 slices representing one 4D volume over one respiratory cycle is selected from the 2800 2D free-breathing dMRI image slices using an optical flux-based optimization method to represent the dynamic thorax of the subject [2]. After 4D construction, we had ∼380 3D images of near-normal subjects and 531 3D images of TIS patients at multiple respiratory phases which included end-inspiration (EI) and end-expiration (EE) respiratory phases.

#### 3. Image intensity standardization

MRI signal intensities in the 4D constructed image are standardized [16] to a standard intensity scale to facilitate MRI segmentation and analysis. Intensity standardization enables voxel intensity values to have similar numeric meaning for each type of tissue within the same subject, across subjects, in repeat scans on the same scanner, and across different scanners [17]

#### 4. Creating ground truth segmentations

Following the principles outlined in [18], we created clinically meaningful and computationally feasible definitions of the thoraco-abdominal body region and objects (Dph, left and right lungs, and thoraco-abdominal skin) in this application to make the models anatomically specific and to minimize inter-tracer variability during creation of the ground truth segmentations of the objects. We define the thoraco-abdominal body region considered in this application as extending from 15 mm superior to the lung apices to the inferior aspect of the kidneys. Similarly, each object was defined in terms of the substructures to be included/excluded. A board-certified radiologist with more than twenty-five years of experience (Torigian) in thoracoabdominal radiology and clinical MRI trained students, engineers, and medical interns (Kogan, Tong L, Mannikeri, Wu, Hosseini, Al-Noury, and Chodvadiya) for anatomic and dMRI radiologic appearance of the relevant structures. Following training, the organs of interest in the (338=189+149) dMRI acquisitions were all segmented manually by the above individuals through use of the open-source software CAVASS [19] in multiple respiratory phases including the EE and EI time points of the respiratory cycle. This yielded a total of > 2,704 (=338×4×2 (EE and EI)) 3D object samples for our cohort.

### 2.1 Dph-R

Dph-R (refer to Figure 2) is based on [9], which was originally developed for localizing thin sparse objects like the thoracic esophagus as well as non-sparse objects like the heart in axial CT images. Dph-R is composed of three modules: backbone network, neck network, and head network. Below, we describe our modifications to adapt this network for our task of localizing the diaphragm on dMRI images.

**Figure 2:**
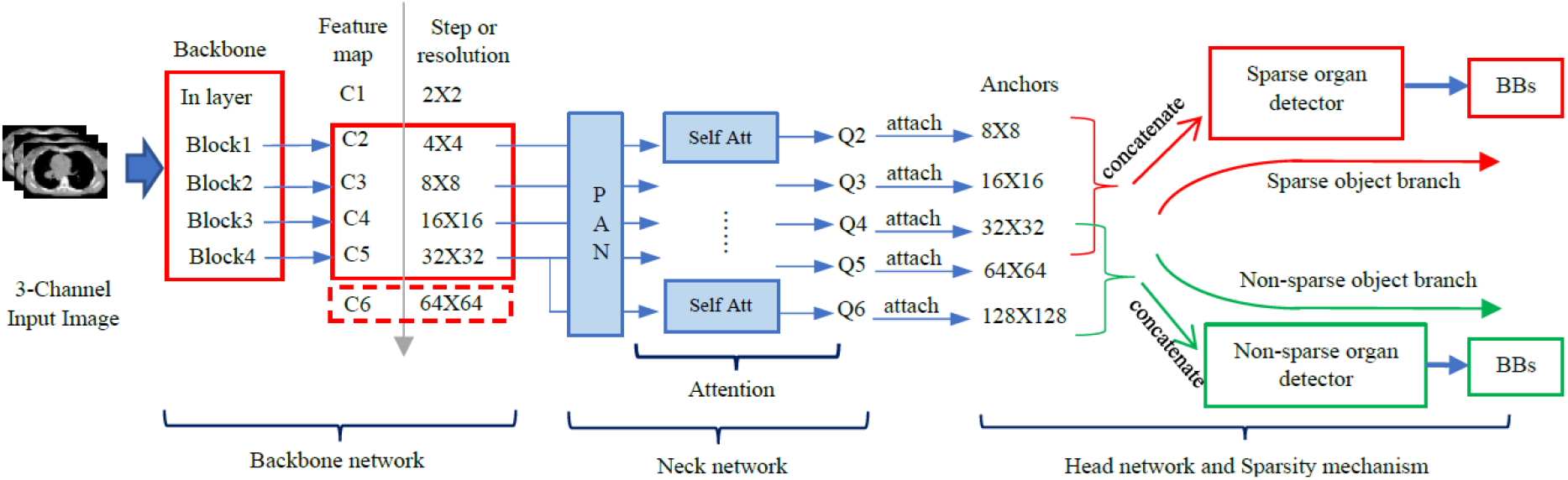
Architecture of DL-R [9] utilized for designing Dph-R.

First, a 2D and 1-channel sagittal slice is converted into a 3-channel image by mapping pixel intensity to a number in each of three pre-defined intensity intervals. This 3-channel slice is taken as input to the backbone network. The weights in the backbone network are initialized from pre-trained models of ResNet [20] and DenseNet [21]. Four feature maps (C2, C3, C4, and C5) are taken as input to the neck network, from the last four convolutional layers of the backbone network using strides of 4, 8, 16, and 32 pixels, respectively. As we move from C2 to C5, the level of information from the 3-channel input image is captured at a more global level.

The neck network utilizes the Path Aggregation Network [10] to merge feature maps (C2, C3, C4, and C5) into maps referred to as Q4, Q5, and Q6 using bottom-up connections, top-down connections, and lateral connections, whereas the Dual Attention Network [11] is used to create prediction maps which have the information dependency across the spatial dimensions and the channel dimensions of the maps Q4, Q5, and Q6. The maps Q4, Q5, and Q6 and the prediction maps are taken as input to the head network.

The head network roughly recognizes the sparse organs with the maps Q4, Q5, and Q6 by associating them with anchor sizes 8 × 8, 16 × 16, and 32 × 32, respectively. The recognition is further refined with the help of convolutional layers by extracting high-level semantic information from the prediction maps. The output from the head network consists of the bounding boxes which localize the object of interest in a sagittal slice. The Dph-R network is optimized using Adam’s optimizer with a learning rate of 0.00001.

### 2.2 Alternate approaches to obtaining bounding boxes for the Dph-D step

We have observed that a Dph-R model created only for the diaphragm does not localize the Dph well in dMRI images. Instead, if a DL-R model is created for 8 organs (Approach_8_organs: two lungs, two kidneys, liver, spleen, skin, and Dph) simultaneously, then the model localizes the Dph well. In this case, it could be that DL-R is able to utilize the additional knowledge about the anatomical layout of different objects to its advantage for localizing the sparse Dph.

The drawback of Approach_8_organs is that the ground truth for 7 other organs besides the Dph are required for creating that DL-R model. The creation of ground truth is labor-intensive and cumbersome, and hence this approach is not practical. Instead, we tried to get the bounding box for the Dph-D step from the auto-segmentations of three other organs (left lung, right lung, and thoraco-abdominal skin) in two ways described below.

#### 2.2.1 Approach_BBox_lungs_skin_2_df

In this approach (refer to Figure 3), the goal is to obtain bounding boxes for the Dph, which are specific to every slice in an image. To do this, we find the inferior most pixel (*p*_*L*_) of an auto-segmentation of a lung in a slice (*s*) if the auto-segmentation is available in *s*. To find the superior (*b*_*s*_) boundary and the inferior (*b*_*i*_) boundary of the bounding box for the Dph in *s*, we take the horizontal lines located 40 mm above *p*_*L*_ and 10 mm below *p*_*L*_, respectively. These distance constants are determined empirically by observation such that the bounding box is large enough to contain the diaphragm. To find the anterior (*b*_*a*_) and the posterior (*b*_*p*_) boundaries of the bounding box in *s*, we locate the two positions (*p*_*a*_ and *p*_*p*_) on the auto-segmentation of the skin in *s* which lie on *b*_*i*_. The vertical line passing through *p*_*a*_ on the anterior side determines *b*_*a*_ and similarly the vertical line passing through *p*_*p*_ on the posterior side determines *b*_*p*_.

**Figure 3:**
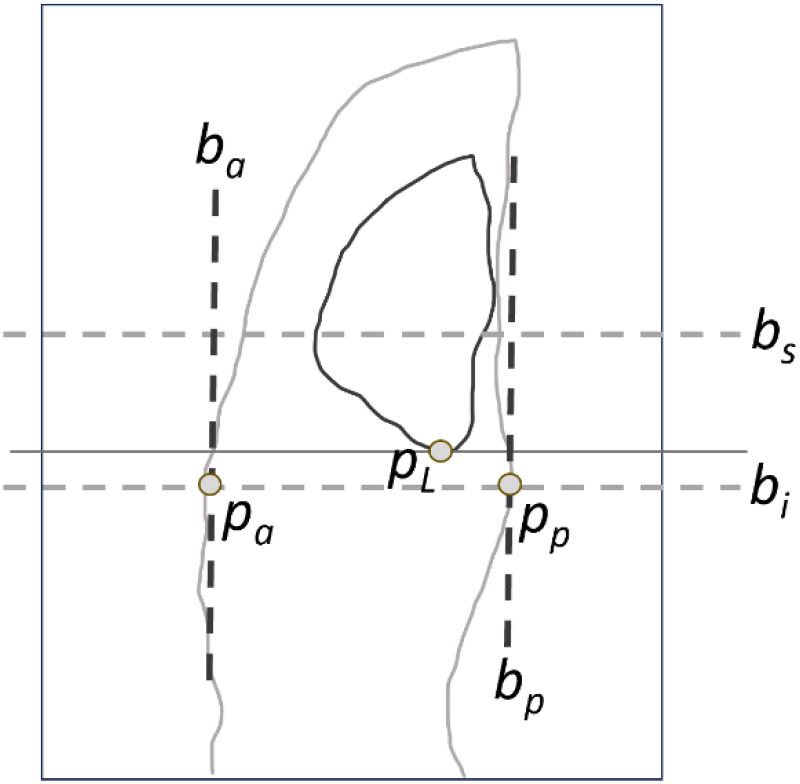
Illustration for obtaining the bounding box for Dph in the sagittal plane through the thorax. The light grey contour and the dark grey contour represent the auto-segmentations of thoraco-abdominal skin and lung, respectively.

The drawback of this approach is that in sagittal locations close to the heart, the lungs are anatomically absent such that no bounding boxes are generated for the diaphragm. Consequently, no diaphragm is segmented in the delineation step on such slices even though the diaphragm is anatomically present. This results in a net poor performance of segmentation for diaphragm on such slices. To overcome this problem, we try the approach shown next.

#### 2.2.2 Approach_BBox_lungs_skin_1_df

In this approach, the idea is to obtain a 3D bounding box for the diaphragm such that the bounding box in a slice is identical in shape and location across all slices. To find this 3D bounding box, we find the smallest 3D bounding box that encloses all the bounding boxes found from Approach_BBox_lungs_skin_2_df. We observe that this approach works well in enclosing the diaphragm across all slices. This approach, despite coarsely localizing the diaphragm, does not compromise the performance of Dph-D in the delineation step. The term ‘df’ in the name of the approaches “Approach_BBox_lungs_skin_2_df” and “Approach_BBox_lungs_skin_1_df” stands for “degrees of freedom” since in the former approach, both the superior-inferior and posterior-anterior boundaries (of the bounding boxes) are variable across slices in an acquisition whereas in the latter approach these two pairs of boundaries are fixed for all the slices in an acquisition.

### 2.3 Dph-D

This module utilizes an encoder-decoder architecture, ABCNet [12], which was originally designed to delineate subcutaneous adipose tissue, visceral adipose tissue, skeletal muscle tissue, and skeletal tissue from low-dose axial CT images of the body torso.

All units of ABCNet are derivatives of BasicConv, which is comprised of four modules in the following order: concatenation, batch normalization, activation, and convolution. There are four DenseBlocks [21] used in the encoder-decoder architecture of ABCNet where each DenseBlock of ABCNet is composed of Dense Layers, which are themselves composed of Bottleneck (a BasicConv with kernel size of 1×1×1) and a BasicConv with a kernel size of 3×3×3 in succession. The lower size of the bottleneck kernel keeps the number of parameters low and simultaneously acts as a feature extractor through the normalization and activation functions of its BasicConv architecture.

ABCNet uses a Dice coefficient-based loss function for training its model and uses patch-based training. The patches are randomly selected from the images in the seen dataset. The output of ABCNet are the prediction maps of the objects of interest in the unseen image, as obtained from the decoder of ABCNet. Using a threshold, these prediction maps are binarized to create the outline of the object of interest. Unlike existing encoder-decoder architectures (DeepMedic, Dense V-Net, V-Net, and 3D U-Net) which have typically 12 or 31 layers and 1 million or 80 million parameters, ABCNet has 118 layers with only 1.4 million parameters. ABCNet is thus attractive because of its deeper architecture with a fewer number of parameters.

### 2.3 Dph-S

The Dph-S utilizes GoogleNet [22] to identify slices which indicate the boundaries of specific body regions. An RNN, as in a bi-directional Long Short-Term Memory (LSTM) module [23], is utilized to improve upon the predictions of GoogleNet. Specifically, the sequence of slices is taken as analogous to a sequence of video frames. The optimum size of the window in the LSTM module of the RNN for recognition of the mid-sagittal plane turned out to be 15+1+15=31 slices. For each of those 31 slices, the 1024-dimensional feature vector of the last max-pooling layer of GoogleNet is taken as input by the RNN.

## 3. RESULTS AND DISCUSSION

The distribution of images for training and testing of DL-R, Dph-R, and Dph-D in all our experiments are shown in Table 0. We show the performance of Dph-R and Dph-D in dMRI images of near-normal subjects in the second and third columns, respectively, of Table 1 where the training and testing of Dph-R and Dph-D have been done with images of 100 and 89 near-normal subjects, respectively, at EE and EI. For Approach_8_organs, Approach_BBox_lungs_skin_2_df, and Approach_BBox_lungs_skin_1_df, the segmentation models (DL-R and DL-D [24]) of objects other than the Dph are obtained with the aforementioned (200 = 100 × 2 (EI and EE)) images.

**Table 0:** Distribution of images for training and testing in experiments of subsequent tables. EE=end expiration.EI=end inspiration.

| Table No. | Module | Training | Testing |
| --- | --- | --- | --- |
| 1 | Approach_8_organs | 200=100 x 2 (EE and EI) near-normal | 178=89 x 2 (EE and EI) near-normal |
|  | Approach_BBox_lungs_skin_2_df |  |  |
|  | Approach_BBox_lungs_skin_1_df |  |  |
|  | Dph-D |  |  |
| 2 | Approach_BBox_lungs_skin_1_df | 378=(100+89) x 2 (EE and EI) near-normal | 100 TIS |
|  | Dph-D | 431 TIS |  |
| 3 | Dph-S | 431 TIS | 100 TIS |
| 4 | Final result of Dph-R, Dph-D, and Dph-S | - | 100 TIS |
| 5 | Models in Table 2 | 378 near-normal and 431 TIS | 20 near-normal and 20 TIS |
| 6 | Models in Table 2 | 378 near-normal and 431 TIS | 20 near-normal and 20 TIS |

**Table 1:** Performance of recognition (location error) and delineation (mean-HD) of the whole diaphragm in 178 dMRI images of near-normal subjects. SD = standard deviation.

| Method | Average±SD location error (mm) | Average±SD mean-*HD* (mm) |
| --- | --- | --- |
| Approach_8_organs | 7.50±7.47 | 1.50±2.32 |
| Approach_BBox_lungs_skin_2_df | 24.32±4.93 | 18.81±18.82 |
| **Approach_BBox_lungs_skin_1_df** | 24.81±5.67 | 1.50±2.21 |

For Dph-R, the performance is reported in terms of location error in units of mm for the whole diaphragm. For Dph-D, we report the results in terms of the average mean-Hausdorff Distance (mean-*HD*) and its SD over all tested samples in the third column. The Dice coefficient (DC) is not an appropriate metric for evaluating the delineation of the Dph since the ratio of the number of voxels in the boundary of the Dph to its volume is ∼1 unlike for compact and large objects for which the ratio is <<1. As such, the DC can be quite low even if the predicted Dph and the true Dph are offset by a small displacement. This problem does not arise with the mean-*HD* metric. The mean-*HD* shown in Table 1 is about one and a half pixels for both the Approach_8_organs and Approach_BBox_lungs_skin_1_df, which shows that the latter approach is a reasonable choice for implementing Dph-R as the disadvantage of the former approach is that it requires the ground truth segmentations of seven other objects other than the Dph.

Utilizing Approach_BBox_lungs_skin_1_df as the Dph-R approach, we report the performance of Dph-D in Table 2 for TIS patients in terms of mean-*HD*. For this experiment, the segmentation models (DL-R and DL-D) for the lungs and the skin were obtained with the aforesaid (378 = (100+89) x 2 (EE and EI)) images of near-normal subjects. For obtaining the Dph-D models, only 431 images of TIS patients have been utilized. The Dph-D model has been tested on 100 different images of TIS patients. The mean-*HD* is about two and a half pixels, which is excellent given that the test dMRI images of Table 2 contain a highly distorted and variable anatomy of the thorax of the TIS patients.

**Table 2:** Performance of delineation of the whole diaphragm in dMRI images of 100 TIS subjects. SD = standard deviation.

|  |  |
| --- | --- |
| **Method** | **Average±SD mean-*HD* distance (mm)** |
| **Approach_BBox_lungs_skin_1_df** | 2.80±2.79 |

Table 3 presents the performance of Dph-S in terms of positional error in units of number of slices and positional error in units of mm in the second and third columns, respectively. For obtaining the Dph-S model, the same 431 and 100 images have been used for training and testing, respectively. We explored three simple variations of Dph-S. First, Google-Net without the RNN (Simple CNN); second, Google-Net without the RNN but using the two neighboring slices as additional inputs to Google-Net (CNN with neighbors); and third, the Google-Net with the RNN (CNN and RNN). We notice from Table 3 that “CNN with neighbors” and “CNN and RNN” perform similarly which motivated us to use the former approach because of its simpler architecture.

**Table 3:**
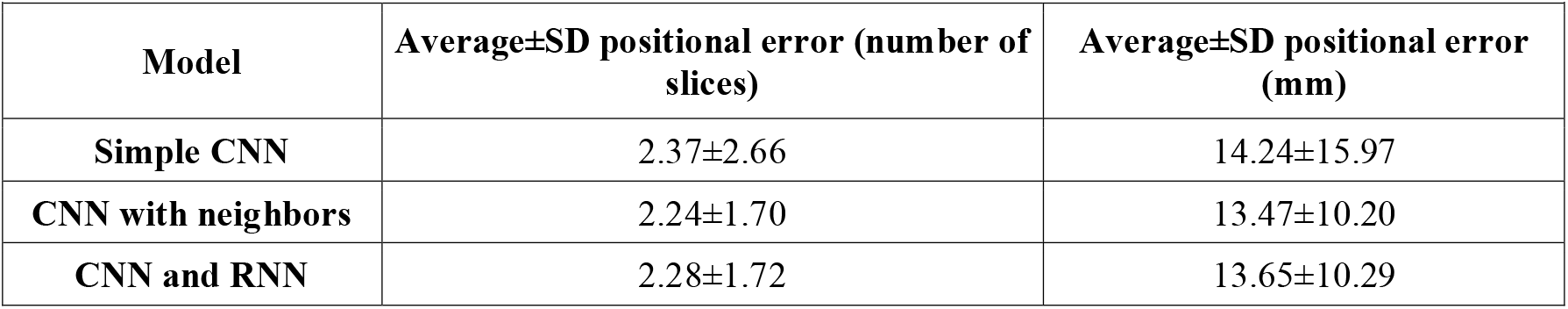
Positional error in number of slices and in units of mm for identifying the mid-sagittal plane in dMRI images of 100 TIS subjects. SD=standard deviation.

|  |  |  |
| --- | --- | --- |
| **Model** | **Average±SD positional error (number of slices)** | **Average±SD positional error (mm)** |
| **Simple CNN** | 2.37±2.66 | 14.24±15.97 |
| **CNN with neighbors** | 2.24±1.70 | 13.47±10.20 |
| **CNN and RNN** | 2.28±1.72 | 13.65±10.29 |

The mean-*HD* in identifying the LHD and RHD by utilizing the prediction of the mid-sagittal plane by “CNN with neighbors” to split the Dph obtained in Table 2 into two parts, LHD and RHD, are shown in Table 4. The mean-*HD* of Table 4 is slightly higher than that in Table 2 since the positional error in identifying the mid-sagittal plane creates an imperfect splitting of Dph into LHD and RHD.

**Table 4:** Performance for segmentation of the left hemi-diaphragm and right hemi-diaphragm in dMRI images of 100 TIS subjects. SD = standard deviation.

|  |  |
| --- | --- |
| **Object of interest** | **Average±SD mean-*HD* (mm)** |
| **Left hemi-diaphragm (LHD)** | 4.24±3.05 |
| **Right hemi-diaphragm (RHD)** | 3.99±3.82 |

We also performed an indistinguishability test to show that our auto-segmentation is indistinguishable from the manual segmentation of the diaphragm. For this experiment, we obtained segmentations of the diaphragm by two other experts (anonymized as Tracer 1 and Tracer 2) who performed the segmentations independently in images of 20 near-normal subjects and images of 20 TIS patients. The mean-*HD* between the auto-segmentation and the segmentations by Tracer 1 and Tracer 2 are shown in the third and fourth columns, respectively, of Table 5. The mean-*HD* between the segmentations by Tracer 1 and Tracer 2 are shown in the fifth column. Paired t-tests indicate that our auto-segmentations are indistinguishable from manual segmentations of the diaphragm when done for images of near-normal subjects.

**Table 5:** Test of indistinguishability between auto-segmentation and manual segmentation (by two anonymized tracers: Tracer 1 and Tracer 2) of the diaphragm in 20 dMRI images of TIS patients and 20 dMRI images of near-normal subjects. SD=standard deviation. EI=end inspiration.

| Datasets | Mean-*HD* (mm) | Between Auto-segmentation and Tracer 1 | Between Auto-segmentation and Tracer 2 | Between Tracer 1 and Tracer 2 | *p*-value ($\alpha=0.05$) |
| --- | --- | --- | --- | --- | --- |
| 20 EI dMRI in TIS patients | Average±SD | 15.88±10.71 | 15.85±10.99 | 1.06±0.37 | <0.0001 |
| 20 EI dMRI in near-normal subjects | Average±SD | 1.11±0.78 | 1.10±0.75 | 0.64±0.15 | 0.08 |

An indistinguishability test for identification of mid-sagittal plane amongst two experts (anonymized as Expert 1 and Expert 2) and by “CNN with neighbors” of Table 3 on the same images used in Table 5, revealed that our Dph-S was comparable to expert identification of the mid-sagittal plane in images of near-normal subjects. The related *p*-values are shown in Table 6. The 3D rendered auto-segmentations and the manual segmentations of the Dph in 6 images are shown in Figure 4.

**Table 6:** P-values for the test of indistinguishability between algorithmic identification and manual identification (by two anonymized experts: Expert 1 and Expert 2) of mid-sagittal plane in 20 dMRI images of TIS patients and 20 dMRI images of near-normal subjects. EI=end inspiration.

| Datasets | Between Algorithm and Expert 1 | Between Algorithm and Expert 2 | Between Expert 1 and Expert 2 |
| --- | --- | --- | --- |
| 20 EI dMRI in TIS patients | 0.002 | 0.001 | 0.91 |
| 20 EI dMRI in near-normal subjects | 0.52 | 0.96 | 0.02 |

**Figure 4:**
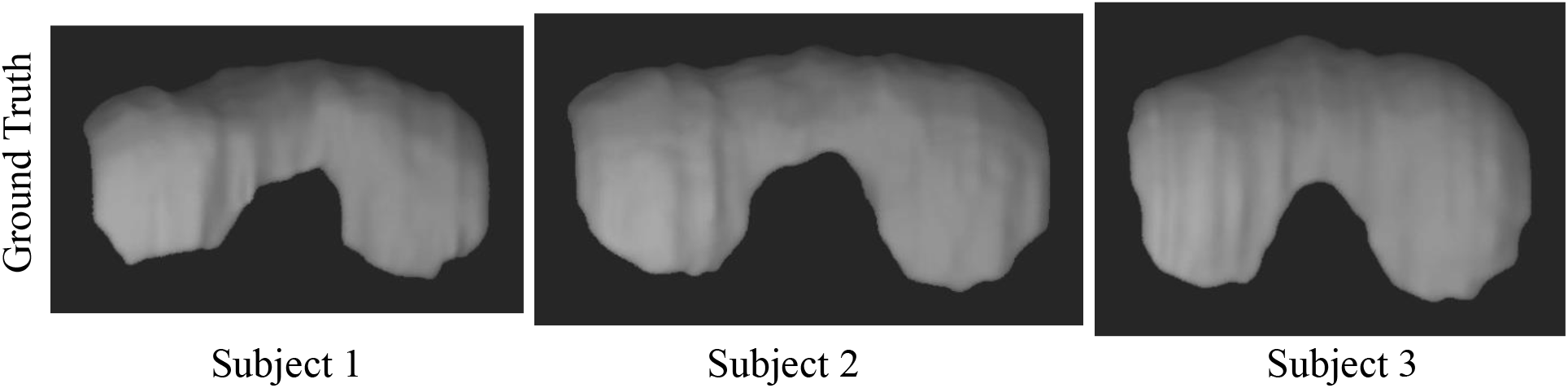
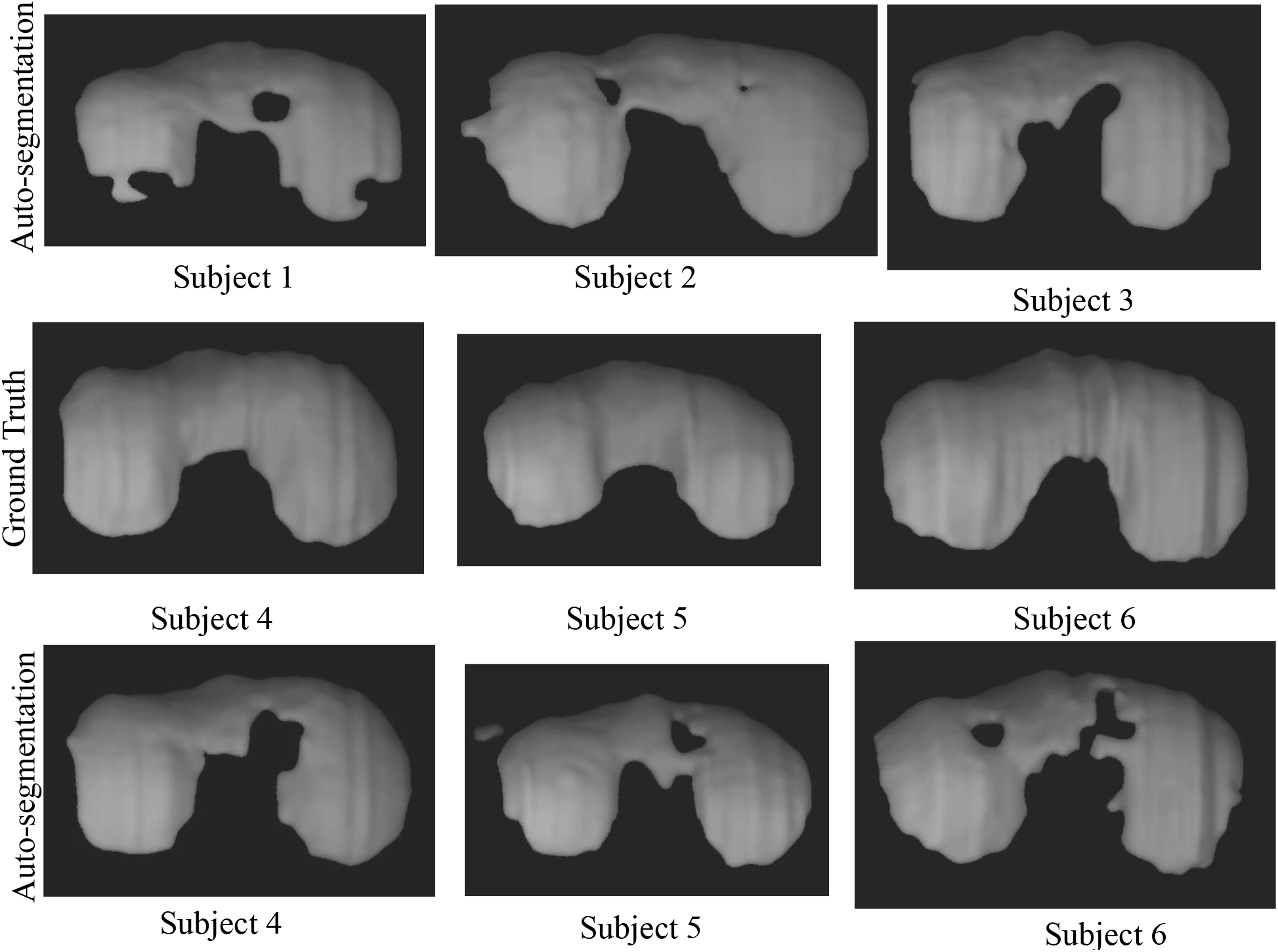
3D rendering of manual segmentations and auto-segmentations of the diaphragm in dMRI images of 6 near-normal subjects. The (deidentified) subject number is listed below the corresponding image.

## 4. CONCLUSIONS

We present a novel and unique system for the challenging problem of segmentation of the diaphragm in dynamic MRI (dMRI) acquisitions of the thoraco-abdominal region, which has not been addressed previously in the literature. This has numerous applications in pediatric and adult respiratory medicine. The mean-*HD* values reported on 100 test dMRI images of TIS patients indicate that our method works reasonably well in segmenting the diaphragm. We have observed that automatic delineations of the diaphragm with our segmentation set-up are indistinguishable from expert annotations of the diaphragm on dMRI of normal pediatric subjects. The results for the delineation of the diaphragm on dMRI of TIS patients are also excellent despite the extreme challenges posed by these images such as the highly deformed and inconsistent anatomy in TIS patients. In future work, we expect to minimize the time required to create ground truth segmentations of the diaphragm in newly acquired dMRI images by refining auto-segmentations of the diaphragm, which would then be used for both building and testing our related models in the process of checking the efficacy of our approach to improve the pre-treatment planning and post-operative assessment of patients with TIS and other disorders.

## Data Availability

All data produced in the present study are available upon reasonable request to the authors.

https://doi.org/10.5061/dryad.vmcvdnczf

## Acknowledgement

This research is supported by a National Institutes of Health Grant: R01HL150147.

